# Is there reporting and citation bias for completed phase III interventional brain tumor trials?

**DOI:** 10.1101/2020.08.10.20172296

**Authors:** Appaji Rayi, Iyad Alnahhas, Pierre Giglio, Vinay Puduvalli

## Abstract

**Aims:** Reporting and citation bias based on positive versus negative outcome among completed phase III interventional brain tumor trials (IBTTs) have not been previously reported. Thus, we aimed to assess the evidence.

**Methods:** Clinicaltrials.gov was used to obtain all phase III IBTTs completed prior to December 31^st^ 2016. Trials closed due to poor accrual or non-phase III were excluded. Data about the funding source, type of intervention, conducted at U.S or Non-U.S locations, patients enrolled, primary completion date, time to dissemination of results in months (either reported on Clinicaltrials.gov or published in a journal), citations per year (from web of science) and outcome (positive or neutral/negative) were abstracted. Median time to dissemination was estimated using Kaplan-Meier estimates and a log rank test for statistical significance. The number of citations per year for positive and negative/neutral studies was compared using a t-test.

**Results:** 67 studies were analyzed. The median time from primary completion to dissemination of results for trials with a positive outcome was 20 months (n = 20; 95% CI: 12-31) compared to 30 months for trials with a negative outcome (n=35; 95 % CI: 22 – 37) (p = 0.095). The average number of citations per year for trials with a positive outcome is 62.4 (n = 21; range 1.1 – 614.8) compared to 25.0 for trials with a negative outcome (n=35; range 0.14 - 158.8) (p = 0.213). There was no significant difference in time to dissemination based on the funding source, type of intervention or the location where the trial was conducted.

**Conclusion:** There was no evidence of reporting or citation bias among completed phase III IBTTs. Nevertheless, positive phase III IBTTs were reported more promptly and cited more often compared to negative or neutral trials. These findings might need consideration for risk of bias assessment while conducting systematic reviews.

## Introduction

Several types of biases such as publication bias, reporting bias, selection bias and citation bias are encountered in the conduct of clinical research. Reporting bias is encountered when the dissemination or reporting of results is altered by the direction of the outcome ^1,2^ and citation bias is observed when there is a bias in citing studies based on the outcome of a study. ^3^ Review of previously published studies among phase III cancer clinical trials has demonstrated that the studies with a positive result are reported earlier, published in higher impact journals, and cited more often compared to those with a negative or null result. ^4^ A high number of negative or null result studies are either unpublished or underreported. To change this trend, several journals have resolved to publish null or negative result studies. ^5^ Reporting and citation biases based on the direction of outcome among completed phase III interventional brain tumor trials (IBTTs) have not been previously reported; here we examined the presence of such bias in such potentially practice changing studies.

## Methods

Data from phase III IBTTs that were completed between inception and December 31^st^ 2016 and registered in the Clinicaltrials.gov database were extracted and included for this analysis. Trials closed due to poor accrual or non-phase III trials were excluded. Data retrieved for the analysis included the type of intervention, the sites of trial conduct (U.S or in non-U.S locations), funding source, demographics of patients enrolled, primary completion date, time to dissemination of results (either as reported on clinicaltrials.gov or in a primary manuscript published in a peer-reviewed journal including the trials results), number of citations per year acquired for the primary publication of the trial (obtained from web of science) and type of outcome of the trial (positive or neutral/negative defined a per previous publication). ^4^ Data about the trials that met the applicable clinical trial (ACT) definition based on the food and drug administration (FDA) amendment act (FDAAA 2007) were also retrieved. ^6^ Median time to dissemination was estimated using Kaplan-Meier estimates and a log rank test used for statistical significance. The number of citations per year for positive and negative/neutral studies was compared using a t-test. A significant level of 0.05 was used for all analyses.

## Results

A cohort of 67 eligible trials was identified with the majority being drug intervention trials, conducted in the US, and involving adult patients with glioblastoma (Table 1). Most of these reported a negative or neutral result (36/67, 54%).

**Table 1.** Summary of trial characteristics

| **Variable** | **Number of trials (%)** |
| --- | --- |
| **Type of intervention** |  |
| Drug | 43 (64%) |
| Device | 4 (6%) |
| Biological | 3 (4%) |
| Radiation | 13 (19%) |
| Other | 4 (6%) |
| **Patient age group** |  |
| Adult (18 - 65 years age) | 52 (78%) |
| Pediatric (<18 years age) | 4 (6%) |
| Older adults (Over 65 Years age) | 3 (4%) |
| All | 8 (12%) |
| **Type of condition treated** |  |
| Glioblastoma | 18 (27%) |
| Recurrent Glioblastoma | 5 (7%) |
| Tuberous sclerosis | 2 (3%) |
| Metastatic tumors | 19 (28%) |
| Low Grade Gliomas | 6 (9%) |
| Meningioma | 2 (3%) |
| Pediatric tumors | 7 (10%) |
| Other | 8 (12%) |
| **Study Funding source** |  |
| NIH | 20 (30%) |
| Industry | 21 (31%) |
| Other | 26 (39%) |
| **Study Site\*** |  |
| U.S | 40 (61%) |
| Non U.S | 26 (39%) |
| **Outcome of Trial** |  |
| Positive | 21 (31%) |
| Negative/Neutral | 36 (54%) |
| Missing outcome data trials | 10 (15%) |
| **Enrollment (IQR)** |  |
| Positive trials | 284 (130 - 414) |
| Negative trials | 347 (193 – 540) |
| Missing outcome data trials | 91 (50 – 141) |
| **Reporting requirement per FDAAA 2007** |  |
| Applicable trials | 30 (45%) |
| Non – applicable trials | 37 (55%) |

The median duration from primary study completion to dissemination of trial results with a positive outcome was 20 months (n=20; 95% CI: 12-31) compared to 30 months for trials with a negative outcome (n=35; 95% CI: 22-37) (p=0.095). The average number of citations per year for trials with a positive outcome was 62.4 (n=21; range 1.1 – 614.8) compared to 25.0 for trials with a negative outcome (n=35; range 0.14 – 158.8) (p=0.213). The median time from primary study completion to dissemination of results for applicable clinical trials (ACT) was 24 months (N=29, 95% CI: 11, 31) compared to 47 months for non-FDA mandated trials (N=36, 95% CI: 28, 70) (p=0.0078). There was no significant difference in time to dissemination based on the funding source (NIH, industry or other sources), type of intervention (drug, device, biological or radiation) or the location (US or Non-US) where the trial was conducted.

## Discussion

We have found that there was no statistically significant evidence of reporting or citation bias between positive and negative/neutral phase III IBTTs. Although, the positive studies were disseminated earlier and cited more frequently compared to negative trials. A relatively smaller number of studies included/analyzed in our cohort might have been the reason for an absence of statistical significance, but the result appears to be encouraging overall. Trials meeting the criteria for an applicable clinical trial, which are by definition those trials which are mandated by the FDA to report their results within 12 months of completion or 24 months post-completion after an extension request, were found to be reporting results in a timely and a significantly earlier period compared to the trials that did not meet the ACT criteria. ^6^ The mandatory requirement of registration and reporting of trial results appears to be critical in reducing some of these biases. Various reasons relating to publication bias typically contribute to reporting and citation bias. ^3, 4^ We included trials from a single database however, Clinicaltrials.gov is a large database consisting over a quarter million registered trials, requiring mandatory registration and reporting if meeting certain criteria and thus the likelihood of excluding a significant number of trials was felt to be low to negatively impact our findings.^6^ There might be several secondary publications reporting the findings of a clinical trial, albeit the primary publication is the most crucial in reporting the primary outcome.

## Conclusion

There was no evidence of reporting or citation bias among completed phase III IBTTs. Nevertheless, positive phase III IBTTs were reported more promptly and cited more often compared to negative or neutral trials. These findings might need consideration for risk of bias assessment while conducting systematic reviews.

## Data Availability

Data will be provided upon request

## Acknowledgements

We thank Dr. Rachel Smith for the statistical assistance.

